# PUBLIC TRANSIT RIDERSHIP ANALYSIS DURING THE COVID-19 PANDEMIC

**DOI:** 10.1101/2020.10.25.20219105

**Authors:** Samira Ahangari, Celeste Chavis, Mansoureh Jeihani

## Abstract

This study investigates the effect of the coronavirus (COVID-19) pandemic on public transport ridership in Baltimore and nine other U.S. cities similar to Baltimore, in terms of population and service area, during the first five months of 2020. The analysis is based on ridership numbers, vehicle revenue hours, and vehicles operated in maximum service. A compliance analysis was done between 2020 and 2019, as well as a monthly analysis of 2020 by mode and type of services. In comparison to 2019, the ridership decreases from March, the start of the pandemic, while all ten cities experienced the most decrease in ridership in April.

## INTRODUCTION

COVID-19, an infectious respiratory virus, first appeared in Wuhan, China, and was pronounced a worldwide pandemic by the World Health Organization (WHO) on March 11, 2020. Such viruses are transmitted from person to person directly via skin-to-skin contact or indirectly via infected surfaces [1]. Since the primary source of contagion happens by breathing droplets from coughs and sneezes [2]–[4], the physical distance between noninfected and infected individuals is essential in preventing the spread of the virus.

Such a pandemic significantly affects transportation systems since a primary response worldwide to slow the epidemic’s spread is to restrict people’s movement. The result of movement restriction was that ridership plunged in all cities. To respond to the ridership decrease, several public transport agencies have reduced their service by keeping key routes and cutting all others, changing from a weekday to a weekend schedule. For example, the Washington Metropolitan Area Transit Authority (WMATA) decreased their services by closing 19 out of 91 metro stations and reducing the service frequencies to 3 or 4 trains per hour from 10 trains per hour [5]. Additionally, WMATA requested that all riders cover their faces while using public transit [5]. In response to the social distancing rule proposed by the government [6], public transit providers redesigned their service while attempting to maintain its functionality.

## LITERATURE REVIEW

Nowadays, the entire world is confronting several challenges, including lockdown, movement restrictions, and schools and businesses closure because of the COVID-19 pandemic that was first recorded in Wuhan, China, in December 2019 [7]. After identifying more than 125,300 cases and 4,981 deaths across the world, WHO declared COVID-19 as a pandemic on March 11. As of July 27, total cases worldwide mounted to 16,296,635, and total deaths were reported at 649,662. The United States has the most deaths (146,968), followed by Brazil (87,004), the United Kingdom (45,837), Mexico (43,680), and Italy (35,107) [8]. It is not the first time in history that humans have faced such a hazard to public health. Other examples include the Spanish Flu or the 1918 flu pandemic, polio in the 1950s, Ebola hemorrhagic fever in 2014, the H1N1 influenza virus in 2009, the 2005-2016 Zika fever, and SARS in 2003.

The COVID-19 pandemic has theatrically altered travel behavior worldwide. In this regard, the effect of COVID-19 on mobility was analyzed for New York City and Seattle as the most infected city [9, p. 19]. One of the sectors most affected by COVID-19 is public transport. North American cities’ ridership dropped by upwards of 90% by the end of March 2020 as governments applied quarantine policies [10]. While the movement restriction and social distancing are in order, public transit’s significant effect cannot be avoided. To respond to the reduced demand, along with a surge of COVID-19 cases among its workforce, New York’s Metropolitan Transit Authority cut its service capacity, especially on express lines and non-rush hour services [11]. Such challenges required public transport agencies to restructure their services, reducing service frequency in some areas while boosting it in others, such as those serving hospitals and essential services. A model proposed to help public transport operators redesign their systems in response to pandemic considered the operational, passenger, and costs of social distancing policies [12]. Two online surveys in South Korea revealed 75.4% and 88.7% of 1,000 respondents, respectively, will not use public transit during the COVID-19 pandemic [13].

The adjustments to services, significantly cutting them, disproportionately affect North American cities with lower-income and more vulnerable populations. A study compared changes in service frequency of 30 U.S. and 10 Canadian cities by considering average income levels and a vulnerability index [10]. Public transit plays an essential role during a pandemic; transit provides vital access to goods and services, especially for riders who do not have access to other modes. Yet public transit is considered by some as a likely environment to spread COVID-19. In 2008, a study in the United Kingdom revealed a high risk of influenza infection using public transport, but the result was not statistically significant [14].

The impact analysis of the severe acute respiratory syndrome (SARS) pandemic in 2003 [15]. The result of several surveys in different places, including Switzerland [16], Chile [17], and Sweden [18], also showed a reduction in public transit ridership. Such a decline is related to government-imposed limitations and travelers’ choices. Public transport stations and vehicles have inadequate physical space available, and their surfaces make the spread of the COVID seem more likely. The estimation shows the highest risk of infection among bus and tram drivers in Sweden [19]. Similarly, China demonstrated the evidence of COVID-19 spread on public buses [20]. An analysis of the influences of COVID-19 on daily public transport ridership in the three most populated regions of Sweden during spring 2020 found reduced ridership [21].

To see whether Americans would respond similarly, this paper analyzed the effect of COVID-19 on public transit riders in 10 cities of the U.S. like Baltimore in terms of population and service size. The purpose of the study is to find the percentage of reduction in ridership in comparison with 2019 and the monthly changes in 2020 by considering the mode and type of service.

## METHODOLOGY

Monthly data reported to the National Transit Database (NTD) since January 2002 was downloaded from the Federal Transit Administration (https://www.transit.dot.gov/ntd/ntd-data). The data consist of “Unlinked Passenger Trips (UPT),” “Vehicle Revenue Miles (VRM),” “Vehicle Revenue Hours (VRH),” and “Vehicles Operated in Maximum Service (VOMS),” each of which was reported by mode and type of service. The data consists of more than 20 methods that, after recategorizing, were considered as three modes – bus (motorbus, commuter bus, trolleybus), rail (light rail, commuter rail, heavy rail, streetcar, and hybrid rail), and demand-responsive (demand responsive vehicle and vanpools)– for this study analysis. Types of service that data reported were Directly Operated (D.O.) and Purchased Transportation (P.T.). Directly Operated service is service that is directly provided by a transit property, while Purchased Transportation is service provided by a third party that usually is a private operator.

For this study, first, nine cities similar to Baltimore in population and size of service area were selected, including Atlanta, Georgia; Denver-Aurora, Colorado; Detroit, Michigan; Minneapolis-St. Paul, Minnesota-Wisconsin; Phoenix-Mesa, Arizona; Riverside-San Bernardino, California; San Diego, California; Seattle, Washington; and Washington, D.C., where transit lines include portions of Maryland and Virginia (**Table 1**). Then the authors analyzed UPT, VRH, and VOMS of each city based on mode (bus, rail, demand-responsive) and type of services (D.O., P.T.). The compliance analysis compared 2019 and monthly changes of 2020.

**Table 1.** Selected Cities and Their Agencies.

| Urbanized Areas | Agency |
| --- | --- |
| Atlanta, GA | Metropolitan Atlanta Rapid Transit Authority |
|  | Georgia Regional Transportation Authority |
|  | vRide, Inc. - Atlanta |
|  | Buckhead Community Improvement District |
|  | Enterprise Rideshare |
|  | City of Atlanta - Department of Public Works - Transit Division |
|  | Georgia State Road and Tollway Authority |
| Baltimore, MD | Maryland Transit Administration |
|  | Baltimore City Department of Transportation |
| Denver-Aurora, CO | Denver Regional Transportation District |
|  | Denver Regional Council of Governments |
|  | vRide, Inc. - Denver |
| Detroit, MI | Suburban Mobility Authority for Regional Transportation |
|  | City of Detroit Department of Transportation |
|  | Detroit Transportation Corporation |
|  | M-1 Rail |
| Minneapolis-St. Paul, MN | Metro Transit |
|  | University of Minnesota Transit |
| Phoenix-Mesa, AZ | City of Phoenix Public Transit Department dba Valley Metro |
|  | Maricopa County Special Transportation Services |
|  | Regional Public Transportation Authority, dba: Valley Metro |
|  | Regional Public Transportation Authority |
|  | vRide, Inc. - Valley Metro |
|  | Valley Metro Rail, Inc. |
| Riverside- San Bernardino, CA | Riverside Transit Agency |
|  | City of Riverside Special Transportation |
|  | Riverside County Transportation Commission |
|  | Ride-On Montgomery County Transit |
| San Diego, CA | San Diego Metropolitan Transit System |
|  | San Diego Trolley, Inc. |
|  | San Diego Association of Governments |
|  | MTS Contract Services |
|  | County of San Diego Transit System |
| Seattle, WA | King County Department of Transportation - Metro Transit Division |
|  | King County Department of Transportation |
|  | City of Seattle - Seattle Center Monorail Transit |
|  | Senior Services of Snohomish County |
|  | Washington State Ferries |
|  | Central Puget Sound Regional Transit Authority |
|  | King County Ferry District |
| Washington, D.C., MD, VA | Washington Metropolitan Area Transit Authority |
|  | DDOT - Progressive Transportation Services Administration |

## RESULTS

We calculated the percentage in ridership (unlinked passenger trips) from 2019 to 2020 for each of the first five months of the year. For example, the percentage change of ridership in March 2020 is equal to the ridership of March 2020 minus ridership of March 2019 divided by ridership of March 2019. As shown in **Figure 1**, the ridership decreases in March, April, and May in all ten selected cities in response to stay-at-home orders that were in effect in March for these cities. In April, ridership was 62-87% less than 2019 levels.

**Figure 1.**
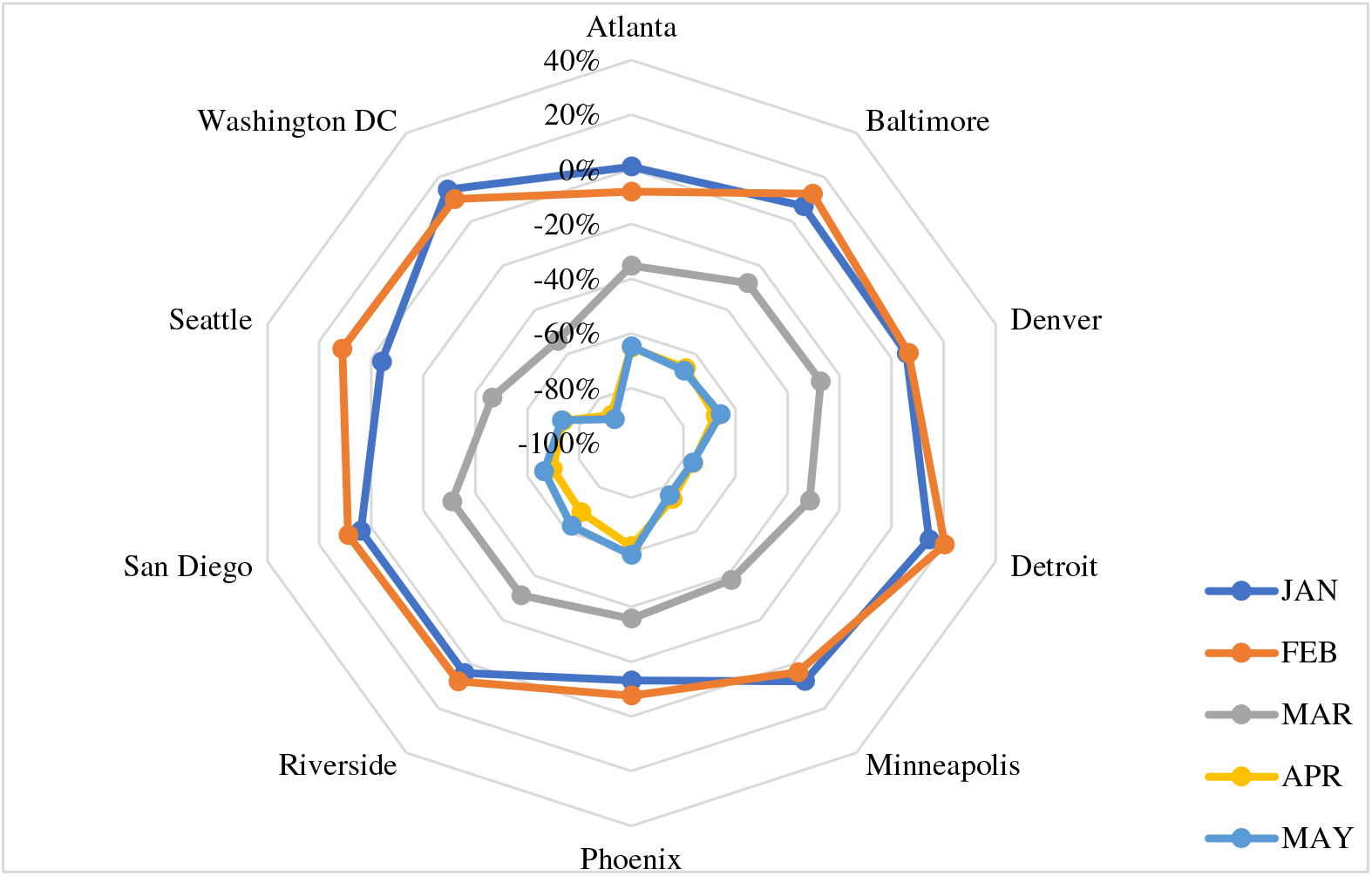
Ridership Change Compared to the Same Month in 2019.

Since COVID restrictions began in March, **Figure 2** compares the change in ridership in each month (Marth through May) compared to February’s base month. As presented in **Figure 2**, ridership started decreasing in March with the lowest ridership in April. Washington, DC, and Seattle saw early declines in ridership. Ridership leveled off in April and May. The cities in California, which were earlier to impose restrictions, saw small gains in May. Washington, D.C., Detroit, and Minneapolis had the most significant ridership drop while the least ridership drop occurred in Phoenix, followed by Atlanta, Baltimore, Denver, San Diego, and Riverside.

**Figure 2.**
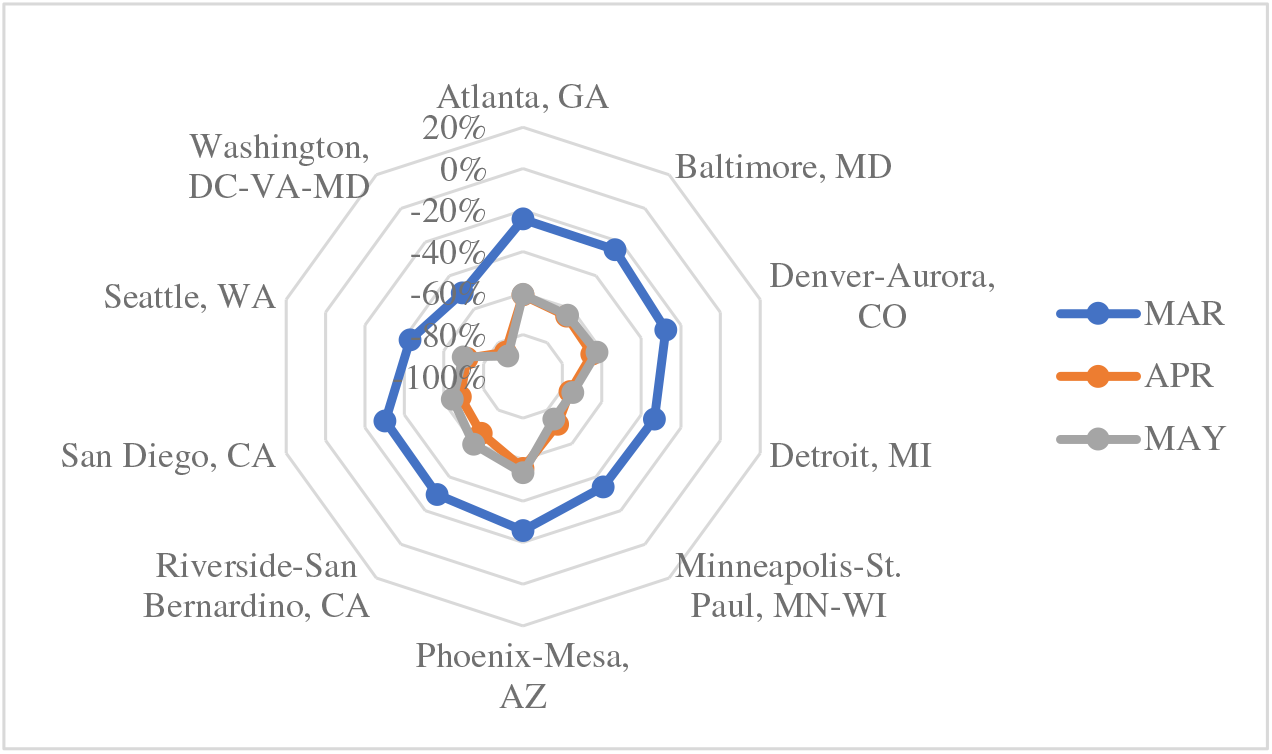
Change in Unlinked Passenger Trips (UPT)

The same result was computed for vehicle revenues hours (VRH), vehicle revenue miles (VRM), and the vehicle operated in maximum service (VOMS). As shown in **Figures 5**, agencies began to reduce service in response to ridership declines. The significant service cuts occurred in April. Service adjustment varied in the ten cities; for example, Detroit had deep cuts in VRH, as did Riverside, Baltimore, and Washington, D.C; see **Figure 3**. Detroit, Baltimore, Washington, and Seattle had steep declines in VRM. As shown in Figure **Figure 4**, agencies took different approaches to VOMS. Seattle only reduces VOMS by 8%, whereas Baltimore and Detroit reduced service by nearly 65% of February service levels. Despite this, the decrease in ridership in Baltimore was slighter than in many of the comparison cities.

**Figure 3.**
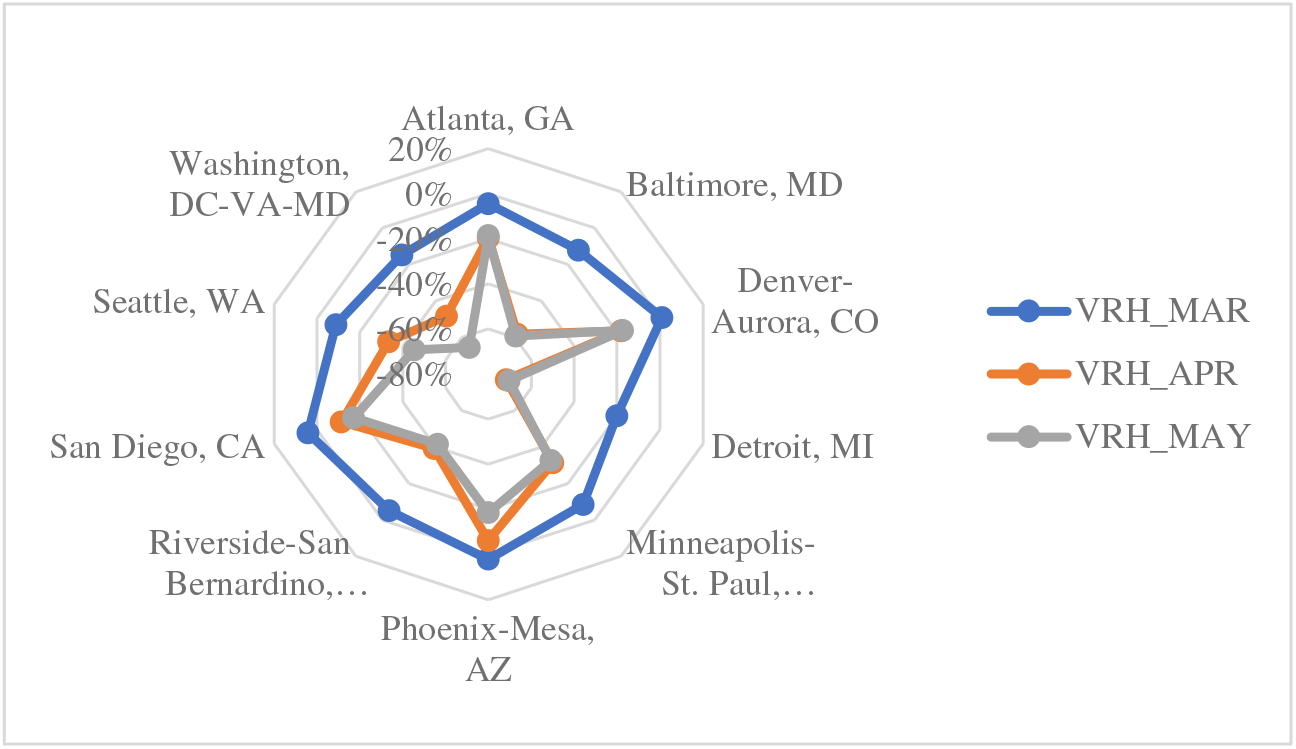
Change in Vehicle Revenue Hours (VRH)

**Figure 3.**
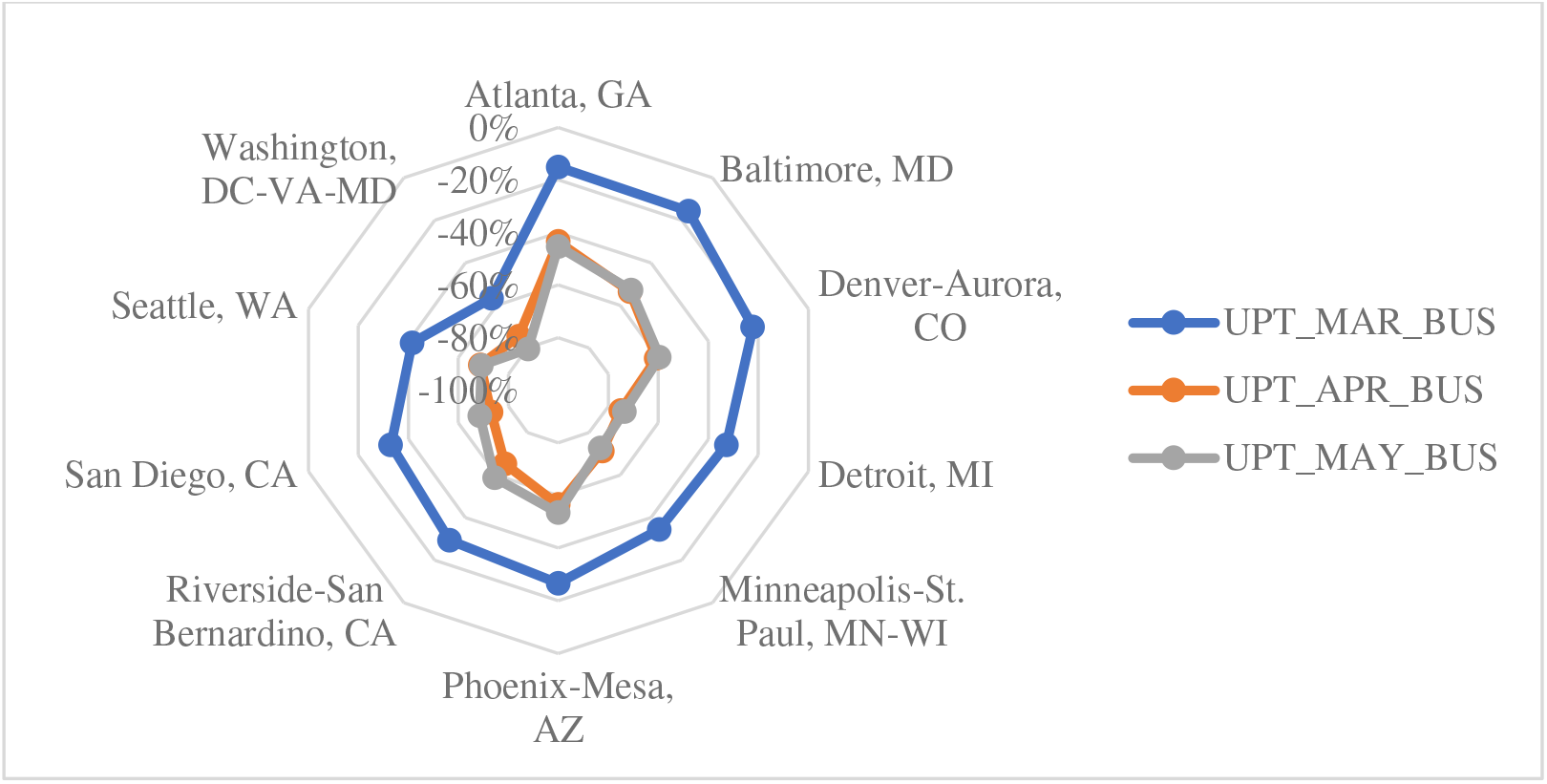
Change in Unlinked Passenger Trips (UPT) for Bus Modes.

**Figure 4.**
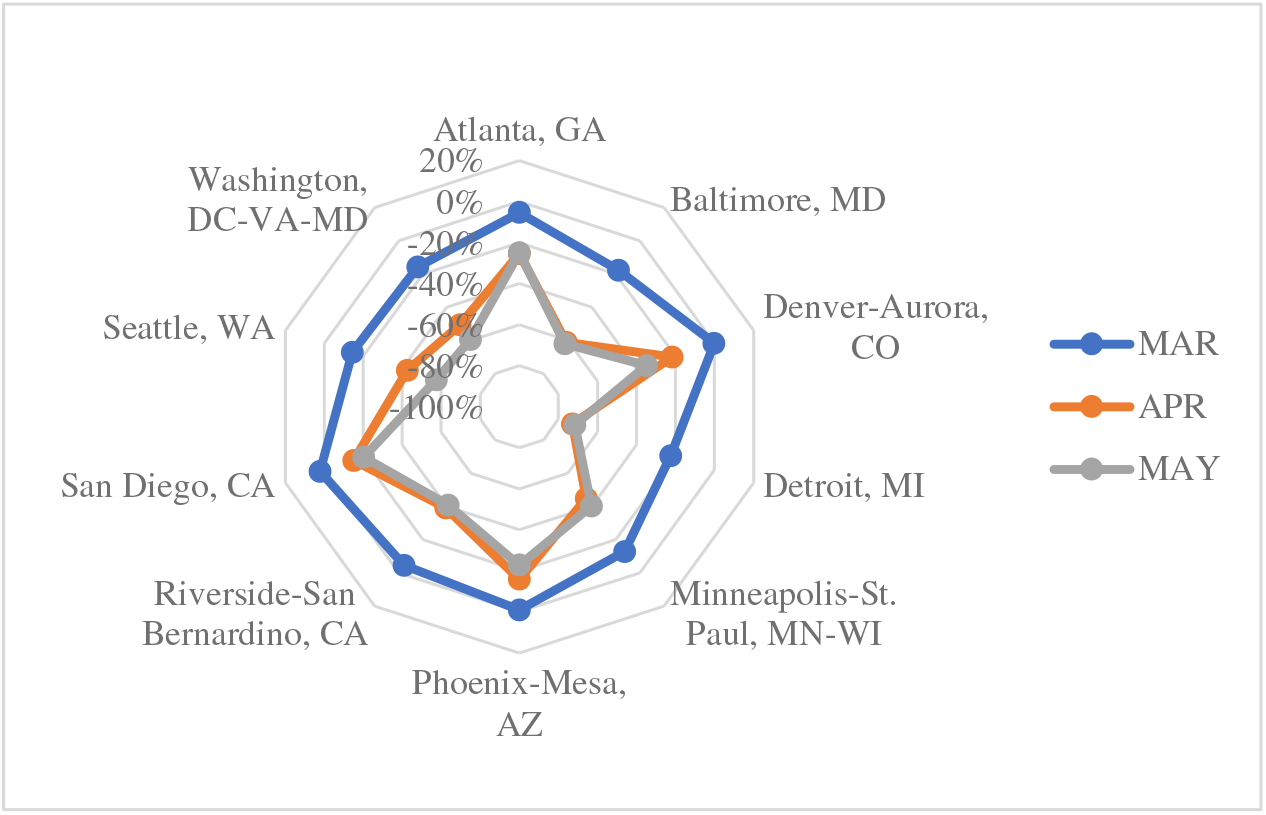
Change in Vehicle Revenue Miles (VRM)

**Figure 4.**
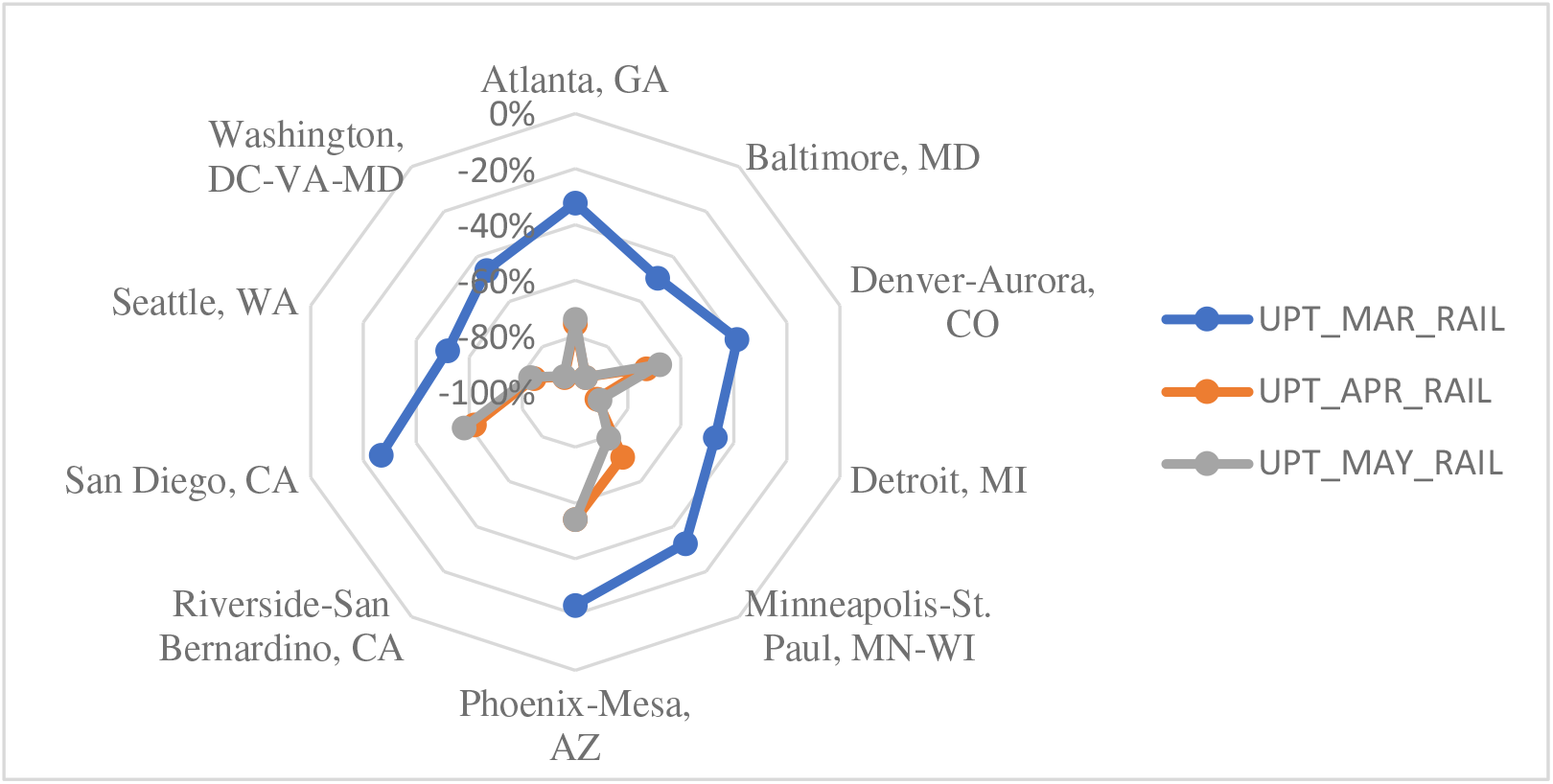
Change in Unlinked Passenger Trips (UPT) for Rail Modes.

**Figure 5.**
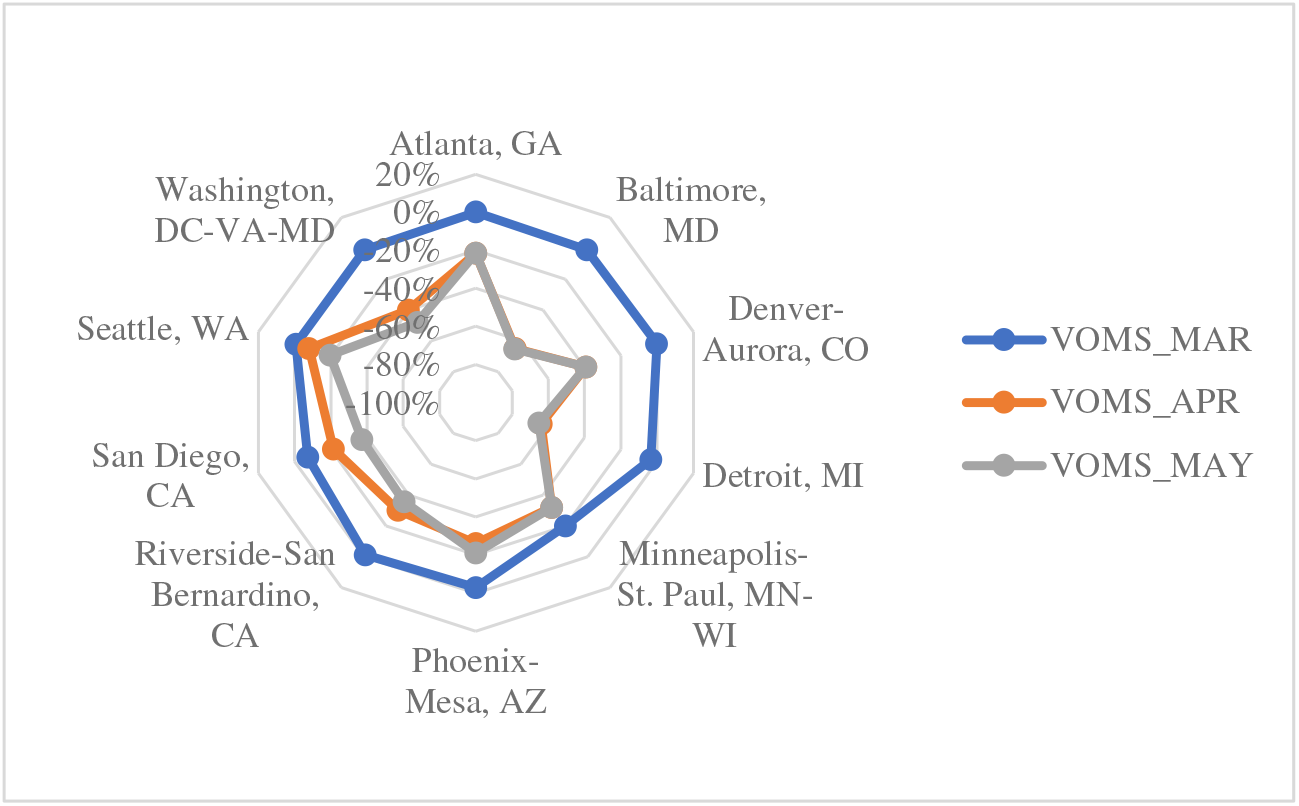
Vehicles Operated in Maximum Service (VOMS)

**Figure 5.**
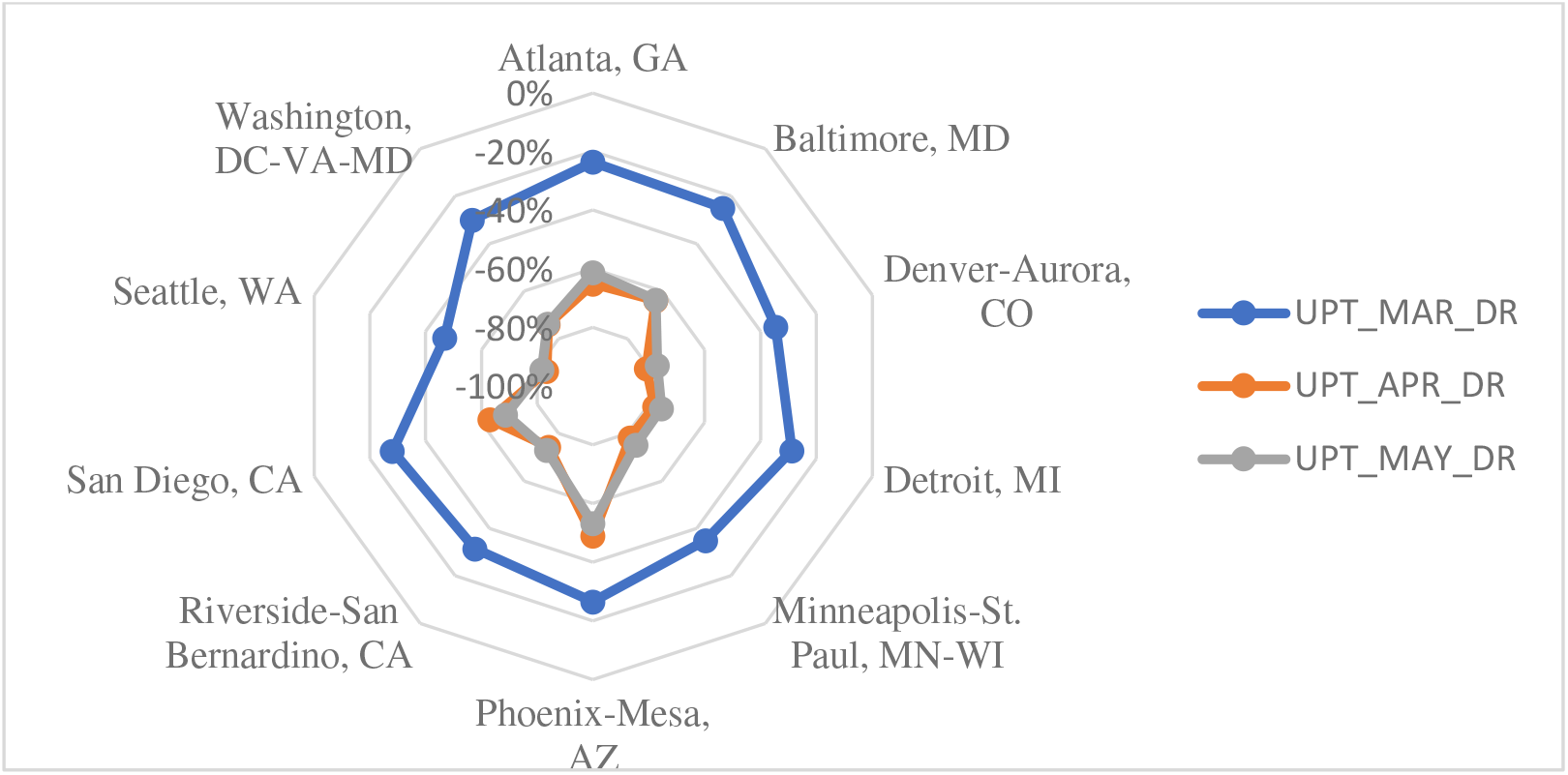
Change in Unlinked Passenger Trips (UPT) for Demand Responsive Modes.

Figure 3 to Figure 5 shows the change in ridership measured by UPTs for bus, rail, and demand-responsive modes. Ridership declines varied by mode. The monthly ridership decreased for all three bus, rail, and demand-responsive modes in March and April compared to February, and ridership remained somewhat steady in May. As shown in Figure 3, Washington, DC, experienced an early decline in bus ridership. By May, the number of UPTs by bus in Washington, DC, was 80% less than in February compared to Atlanta, where bus ridership only declined by 45%. Rail modes generally experienced steeper declines in ridership. Compared to February, UPTs for rail modes declined by 54-93% by May; see Figure 4. Riverside does not have any rail services in operation. Demand responsive trips had the least decline in ridership. As shown in Figure 5, in May, UPTs were 53-82% less than the number of UPTs in February.

## Regression Analysis

To find the factors affecting transit ridership, several regressions analysis was performed. Sociodemographic data for each urbanized area was obtained using the 2018 American Community Survey (ACS) 1-year estimates. The unemployment rate for February through May was obtained from the Bureau of Labor Statistics (BLS) by metropolitan statistical area (MSA). As shown in Figure 6, the unemployment rate increased rapidly in April, with some regions showing a slight recovery in May. The Detroit area was hit very hard by COVID, one in four individuals was unemployed. Lastly, we considered the labor force in each MSA. Using 2019 BLS data, the total number employed in each primary labor industry was found. Based on the job category, we categorized the industry type as essential and non-essential. We considered necessary jobs as jobs that individuals could not do from home. We evaluated the percentage of people working in critical positions and the average hourly and salary wages for essential jobs.

**Figure 6.**
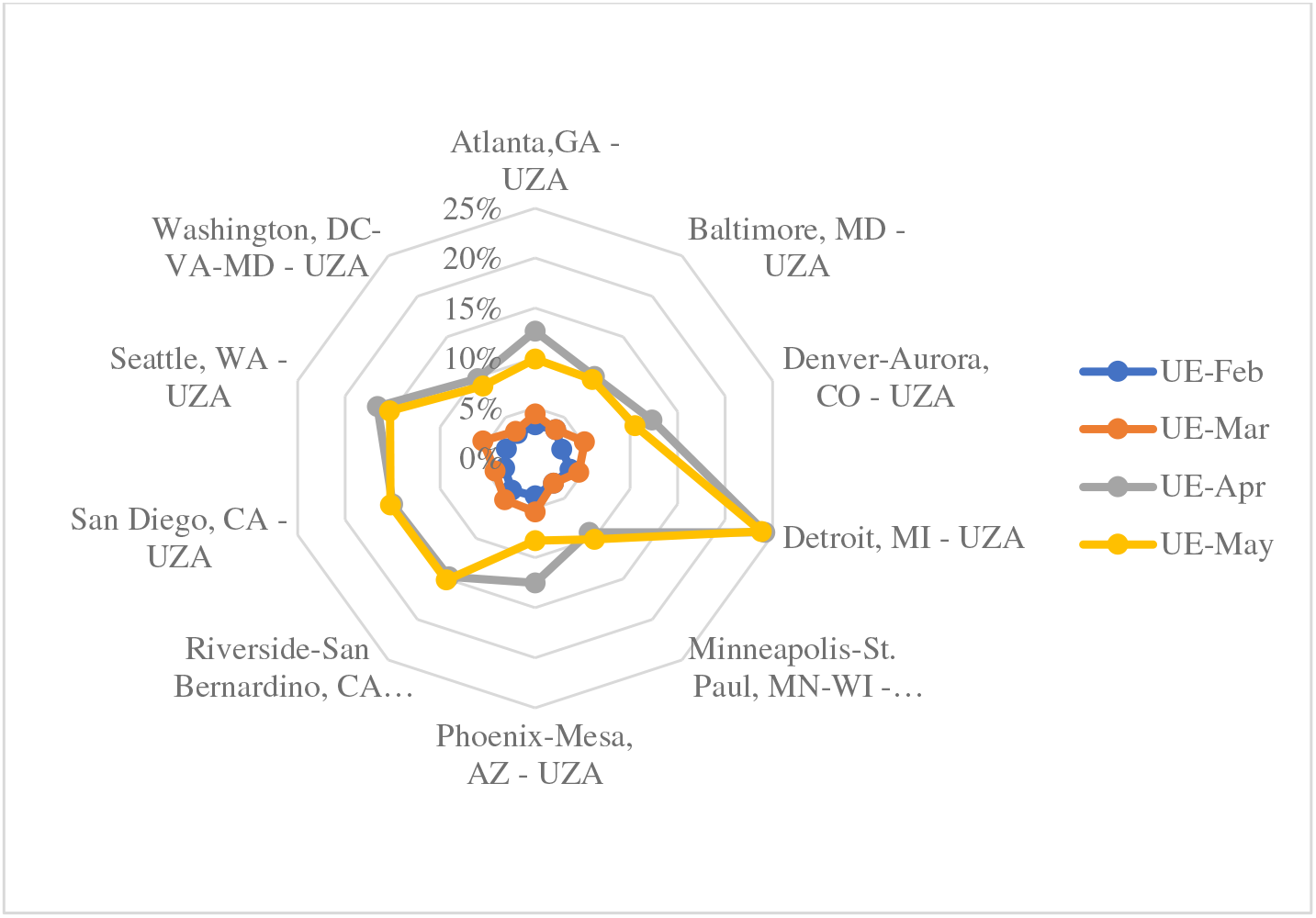
Unemployment Rates Each Month.

**Table 3** presents the results of the regression analysis for predicting the number of unlinked passenger bus trips. The significant factors were the percentage of poverty, percentage with H.S. degree or higher, percentage of foreign-born, the portion of taking public transit to work and do not own a vehicle, and the unemployment rate. As the poverty rate increased, the number of bus trips increased. Conversely, higher unemployment rates reduced the likelihood of trips. The result in **Table 4** shows that the only significant factor is the unemployment rate for trips by rail. The higher the unemployment rate, the more decrease in ridership. The lack of substantial socioeconomic factors highlights the different roles each modality plays; rail tends to serve choice riders. The result of **Table 5** shows that median income, unemployment rate, percentage of essential business labor group, the hourly wage of the crucial group, and vehicle revenue miles for demand responsive transit were significant factors in predicting the percentage change in UPTs. Demand responsive trips were the only modality where an operational variable was substantial. Unlike bus and rail, where service changes may be in response to lower ridership, changes in demand-responsive service directly impact ridership.

**Table 2.** Sociodemographic Data for Select Urbanized Areas.

| Urbanized Area | Median Income | Percent In Poverty | % H.S degree or higher | % Bachelor's degree or higher | % Non-English Speaking | % Foreign Born | % Households No Vehicle | % Take Public Transit to Work | % Take Public Transit that Doesn't Own Vehicles |
| --- | --- | --- | --- | --- | --- | --- | --- | --- | --- |
| Atlanta, GA | 70342 | 10.9% | 90.6% | 42.3% | 20.1% | 15.5% | 3.1% | 3.4% | 35.3% |
| Baltimore, MD | 76111 | 11.1% | 90.7% | 41.3% | 15.0% | 12.3% | 5.4% | 7.3% | 35.6% |
| Denver-Aurora, CO | 76959 | 8.4% | 90.9% | 44.4% | 20.8% | 12.5% | 2.2% | 4.0% | 15.0% |
| Detroit, MI | 58474 | 15.2% | 89.7% | 32.3% | 15.5% | 11.3% | 3.4% | 1.4% | 42.9% |
| Minneapolis-St. Paul | 77580 | 9.2% | 93.6% | 45.7% | 17.7% | 13.0% | 3.5% | 5.4% | 22.2% |
| Phoenix-Mesa, AZ | 63648 | 12.3% | 88.3% | 33.3% | 27.3% | 14.9% | 2.7% | 1.9% | 36.8% |
| Riverside-San Bernardino, CA | 69254 | 13.7% | 78.8% | 20.4% | 49.4% | 23.4% | 1.5% | 1.6% | 18.8% |
| San Diego, CA | 78104 | 11.5% | 87.2% | 38.6% | 38.8% | 24.1% | 2.4% | 2.7% | 22.2% |
| Seattle, WA | 87649 | 9.1% | 9.3% | 45.4% | 25.8% | 20.8% | 4.7% | 11.6% | 16.4% |
| Washington, DC-VA-MD | 103474 | 8.0% | 91.1% | 54.5% | 33.1% | 26.0% | 6.5% | 15.2% | 23.0% |

**Table 3.**
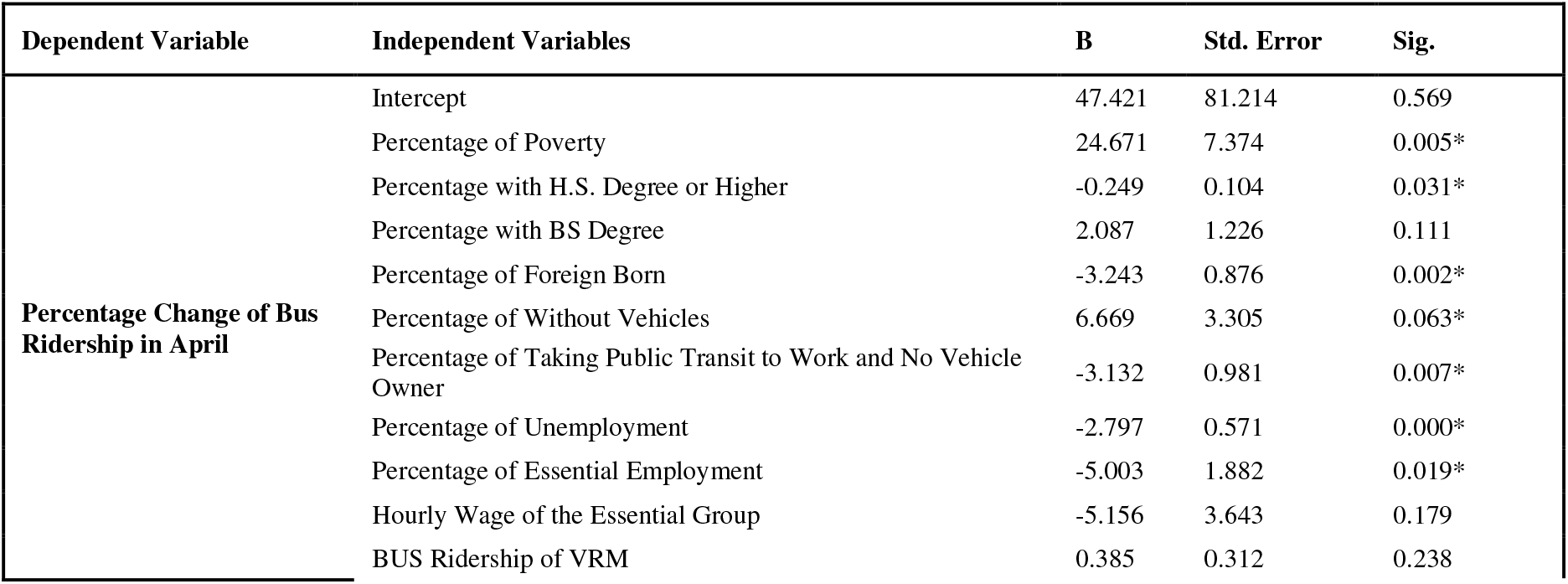
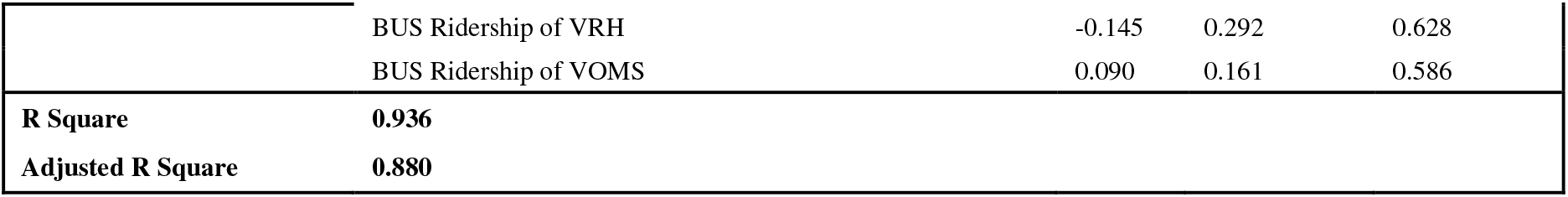
Regression Analysis of Bus Ridership.

**Table 4.** Regression Analysis of Rail Ridership.

| Dependent Variable | Independent Variables | B | Std. Error | Sig. |
| --- | --- | --- | --- | --- |
| **Percentage Change of Rail Ridership in April** | Intercept | -332.623 | 157.932 | 0.053* |
|  | Percentage of Poverty | 2.347 | 5.036 | 0.648 |
|  | Percentage with H.S. Degree or Higher | -0.125 | 0.248 | 0.623 |
|  | Percentage with BS Degree | 3.149 | 2.283 | 0.190 |
|  | Percentage of Non-English Resident | 0.827 | 0.718 | 0.269 |
|  | Percentage of Without Vehicles | -3.337 | 14.140 | 0.817 |
|  | Percentage of Taking Public Transit to Work | -1.182 | 5.734 | 0.840 |
|  | Percentage of Taking Public Transit to Work and No Vehicle Owner | 1.253 | 1.166 | 0.301 |
|  | Percentage of Unemployment | -2.284 | 1.081 | 0.053* |
|  | Percentage of Essential Employment | 4.302 | 2.640 | 0.126 |
|  | Rail Percentage of VRM | 0.012 | 0.433 | 0.978 |
|  | Rail Percentage of VRH | 0.608 | 0.538 | 0.277 |
|  | Rail Percentage of VOMS | -0.289 | 0.194 | 0.159 |
| **R Square** | **0.88** |  |  |  |
| **Adjusted R Square** | **0.784** |  |  |  |

**Table 5.**
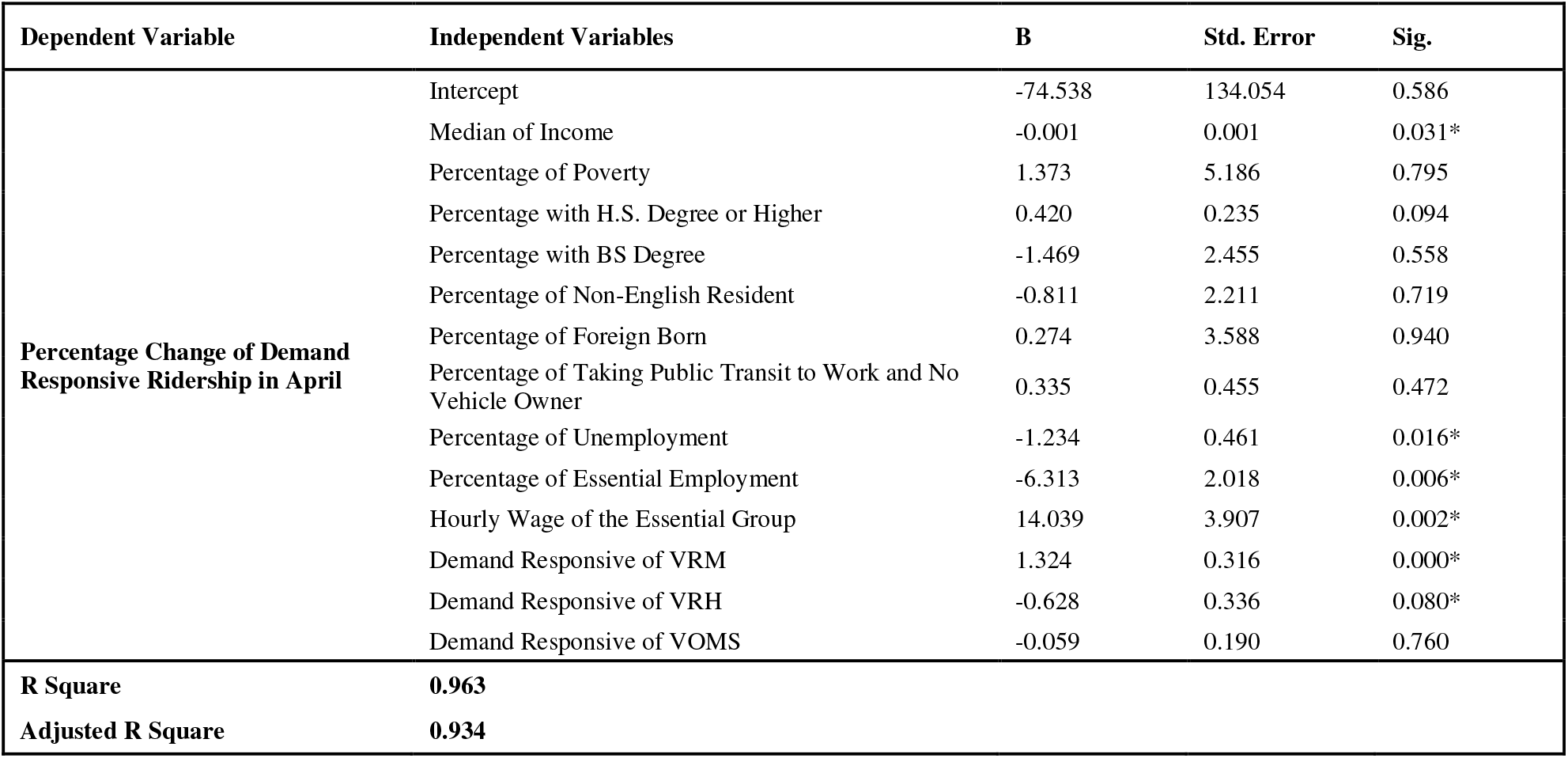
Regression Analysis of Demand Responsive Ridership.

## CONCLUSIONS

The COVID-19 pandemic affected transportation worldwide due to both stay-at-home restrictions and individual riders’ choice to avoid transportation modes where social distancing is difficult, and numerous people touch surfaces. Transit agencies responded by cutting service and hours and redirecting some services to high-need areas such as hospitals. Simultaneously, essential workers still needed transportation to their jobs, and such cuts disproportionately affected more impoverished areas.

The authors chose ten cities for this analysis. After first classifying some 20 modes of transportation into three groups of the bus, rail, and demand-responsive, this study compared ridership and metrics such as vehicle revenue hours, vehicle revenue miles, and vehicles operating in full service against both 2019 numbers and monthly figures for the first five months of 2020.

The results show that ridership, not surprisingly, decreased in March as the virus spread and stay-at-home orders were issued and hit its lowest peak in April in all ten cities.

The regression analysis showed that the only factor affecting rail ridership reduction in April 2020 was the unemployment rate. In demand-responsive mode, income, unemployment rate, the percentage of essential unemployment, and vehicle revenue hours (for demand-responsive mode) directly affected the ridership reduction. In contrast, vehicle revenue mile and the crucial group’s hourly wage had a reverse effect on ridership reduction. In the bus mode, which captures captive users; poverty, education. (percentage with high school degree or higher), the percentage of foreign-born residents, the percentage of residents who take public transit to work, and do not own a vehicle, and the unemployment rate causes a higher reduction in bus ridership in April. The percentage of people without a vehicle yields a lower decrease in ridership.

## Data Availability

https://docs.google.com/forms/d/1qOY4LCLEWmpMtPZYDlE5UmmRqCmUuhIi7-R97EAcPF4/edit#responses

## ACKNOWLEDGMENTS

This study was funded by the Urban Mobility & Equity Center at Morgan State University and the University Transportation Center(s) Program of the U.S. Department of Transportation.

